# Establishment of a teaching hospital-based dementia consultation service for rurally-based regional district general hospitals

**DOI:** 10.1101/2021.10.14.21264940

**Authors:** Björn H. Schott, Jakob Christian Voetlause, Juliana Lisa Amoah, Alexander Kratzenberg, Michael Belz, Tobias Knipper, Charles Timäus, Carmen Beskow, Catherine M. Sweeney-Reed, Jens Wiltfang, Katrin Radenbach

## Abstract

**Objective:** The treatment of patients with dementia poses a considerable challenge to regional district general hospitals, particularly in rural areas. Here we report the establishment and initial evaluation of a dementia-specific consultation service provided by a teaching hospital-based Psychiatry Department to regional district general hospitals in surrounding smaller towns.

**Methods:** The consultation service was provided to patients with pre-existing or newly suspected dementia, who were in acute hospital care for concurrent conditions. An evaluation of 61 consultations – 49 on-site and 12 via telemedicine – was performed to assess the needs of the participating hospitals and the specific nature of the referrals to the consultation service.

**Results:** Suspected dementia or cognitive dysfunction was the primary reason for consultation requests (>50% of cases). Other common requests concerned suspected delirium, behavioral symptoms, and therapeutic recommendations. During the consultations, a diagnosis of dementia was reached in 52.5% of cases, with other common diagnoses including delirium and depression. Recommendations related to pharmacotherapy were given in 54.1% of consultations. Other recommendations included referral for outpatient neurological or psychiatric follow-up, further diagnostic assessment, or assessment in a memory clinic. Geriatric psychiatric inpatient treatment was recommended in only seven cases (11.5 %).

**Conclusions:** Our initial evaluation demonstrates the feasibility of providing a dementia-specific consultation service in rural areas. The service has the potential to reduce acute transfers to inpatient geriatric psychiatry and enables older patients with dementia or delirium to be treated locally by helping and empowering rurally-based regional hospitals to manage these problems and associated complications.

## 1 Introduction

Patients with dementia admitted to hospital for acute treatment of concurrent conditions are at increased risk of complications and adverse events such as falls, delirium, or severe behavioral symptoms, culminating in overall higher mortality (Inouye et al., 2014b; Dewing and Dijk, 2016; Blair et al., 2018). Advanced age is the primary risk factor for developing dementia, and the proportion of individuals affected by dementia is therefore increasing steeply in aging populations, such as in most European countries(Cao et al., 2020).

The situation of people with dementia in hospital is frequently complicated by other related conditions, which may present similarly, yet necessitate a different or more comprehensive treatment approach. A cardinal example is delirium, which is common following surgery (Ní Chróinín et al., 2021), and can be aggravated by inadequate medication (Inouye et al., 2014b). Delirium typically results in prolonged length of inpatient stay and a short-term 20-fold increase in mortality. A quarter of older patients with delirium die within three to four months, and 41% of previously self-caring individuals require discharge to a nursing facility. Symptoms are fully reversible in approximately 50% of cases (Inouye et al., 2014a), but recovery may depend on timely recognition and initiation of appropriate treatment. Another challenge in the care of the elderly, and especially patients with dementia, is the frequent polypharmacy, which refers to the use of four to five or even more medications at the same time. Even in the absence of obvious side-effects, polypharmacy must be regarded as a risk factor for the occurrence of complications like delirium or behavioral disorders. Such complications can arise because numerous pharmaceuticals, for example loop diuretics (Sica, 2004) or digitalis glycosides (Marvanova, 2016), are generally capable of triggering delirium in predisposed patients. Furthermore, psychotropic medications frequently used for behavioral disorders may themselves induce considerable somatic and psychiatric side-effects (Inouye et al., 2014a).

Several approaches have been proposed to improve the care for patients with dementia and to prevent delirium in acute care hospitals. Interventions to improve care in general include patient-centered care provided by hospital staff (Chenoweth et al., 2021) or the training of volunteers (Blair et al., 2018). To specifically reduce the incidence of delirium, both non-pharmacological (Burton et al., 2021) and pharmacological (Park et al., 2021) interventions show moderate effectiveness, but there is considerable variability concerning the benefit for the individual patient.

While these approaches are promising and are currently being further developed, their applicability may be particularly challenging in smaller, rural hospitals: In addition to the increasing average age of the population, younger individuals frequently concentrate in larger cities, leaving small towns and rural areas predominantly affected by the consequences of having an aging population (Milbert and Sturm, 2016). This trend leads to a disproportionately increasing number of patients with dementia in rural hospitals, and at the same time, contributes to the lack of qualified staff to support specialized units for dementia treatment and care, such as neurology or (geriatric) psychiatry departments (van den Berg et al., 2011; Mathur et al., 2019). Here we propose and evaluate the feasibility of an out-reach consultation service provided by a specialized institution within rural surroundings to alleviate this problem.

We report the establishment of a dementia-specific consultation service to improve the care for patients with dementia treated in rurally-based regional hospitals. The service as described here was provided by the Department of Psychiatry and Psychotherapy of the University Medical Center Göttingen, Germany, and was available for patients treated in local regional hospitals upon request by their treating clinicians. An initial evaluation was performed to assess the needs and questions commonly raised by the treating hospitals as well as recommendations commonly made by the providers of the service.

## 2 Methods

### 2.1. Establishment of a dementia consultation service

#### 2.1.1. Preparatory work

The proposal for establishing a dementia-specific consultation service for rurally-based regional hospitals was initially developed as part of an initiative funded by the Department of Social Affairs, Health and Equality of the State of Lower Saxony (2019). The study was approved by the Ethics Committee of the University Medical Center Göttingen (approval number 13/12/19) and was registered with the German Clinical Trials Register (registration number DRKS00022366).

#### 2.1.2. Setting and participating hospitals

The consultation service was provided by the Department of Psychiatry and Psychotherapy of the University Medical Center Göttingen (UMG). The university city of Göttingen, with a population of 118,911 (December 2019, https://www.statistik.niedersachsen.de), is surrounded by the rural counties of Göttingen (county), Goslar, and Northeim. We identified nine hospitals without in-house Psychiatry and/or Neurology Departments in the assigned service area of the Department of Psychiatry and Psychotherapy of the UMG.

#### 2.1.3. Team

The team consisted of two senior residents in psychiatry (T.K. and C.T.), a nurse with long-standing experience in geriatric psychiatry and dementia care (C.B.), as well as a project coordinator (A.K.) and two research students (J.C.V. and J.L.A.). Organizational and clinical supervision was carried out by two senior psychiatrists specialized in geriatric psychiatry and/or consultant liaison psychiatry (K.R. and B.H.S.).

#### 2.1.4. Legal issues and service agreements

The service agreement was drawn up in collaboration with the UMG’s legal department. After a formal presentation of the project was held in a partner hospital expressing interest, the written service agreements were sent to the respective hospital for review if further interest was expressed. Upon mutual agreement, the service agreements were signed by the Medical Director of the Department of Psychiatry and Psychotherapy of the UMG and the board members (UMG and partner hospital). Service agreements were established with five partner hospitals, including four general care hospitals and one hospital specialized in interdisciplinary diabetes care. Agreements were signed between 18.05.2020 and 21.07.2020.

### 2.2. Implementation of the consultation service

#### 2.2.1. Request and performance of consultations

Information regarding the process of requesting consultations, including contact options, was provided to the hospitals as a flowchart during the initial project visit. Consultations could be requested by phone, e-mail, or fax. They included history and neuropsychiatric examination and were performed either on-site or via telemedical service, and were followed by recommendations regarding diagnostic and therapeutic measures. Further diagnostic assessment for the differential diagnosis of a dementia syndrome or its complications was recommended where applicable, as well as specific therapies and support from Social Services. Specific attention was paid to complex polypharmacy, and advice was given by physicians on how to avoid possible pharmacological risk factors for deterioration of the patient’s condition (e.g., by avoiding potentially delirium-inducing substances).

An educational program complemented the consultation service. The program included both scheduled talks on selected topics concerning dementia treatment and care as well as hands-on teaching. A nurse with long-standing experience in geriatric psychiatry and dementia care supported the nursing staff on-site by providing advice on the nursing management of patients with dementia, including means of communication and improving the hospital environment to meet the special needs of these patients. Advice was provided both in general and on a case-by-case basis. A detailed evaluation of the educational program and the local expertise form part of a separate, ongoing, study.

#### 2.2.2. Standardized documentation of consultations

A standardized form was used for systematic clinical documentation of the consultations, including the questions leading to consultation, a detailed medical and psychiatric history, and the recommendations made by the providing physician. The questionnaire was available in both digital and paper form (see Appendix). The Mini-Mental State Examination (MMSE) was additionally administered to a total of 40 patients. While primarily designed to meet the clinical needs of the requesting institutions, these standardized forms also provided the basis for the statistical evaluation of the consultations reported here.

### 2.3. Preliminary evaluation

#### 2.3.1. Data extraction and statistical analysis

A central aspect of the current study was the evaluation of the questions raised by the partner institutions when requesting our consultation service for an individual patient. The circumstances leading to initiation of the consultations were also considered. Since the wording of questions arising under comparable circumstances varied, we performed a content analysis using inductive categorization to enable quantitative analyses (Mayring and Fenzl, 2014). As consultation forms were primarily completed to meet the needs of the referring clinicians, the categories were defined according to the existing content. For example, the category *Suspected Dementia* included questions regarding dementia, memory impairment, and cognitive decline, whereas questions concerning confusion and disorientation were also assigned to the category *Suspected Delirium*. Furthermore, questions relating to drug intolerance and to recommendations on pharmacotherapy were included in a single category (*Pharmacological Issues*): referring clinicians typically requested advice regarding a potential change or discontinuation of medication in both areas.

Given the preliminary nature of the current evaluation, we report our findings primarily as frequency data. Where inference statistics were applicable, *X*^2^ tests were used to evaluate frequency deviations, and t-tests were applied to continuous data. Owing to the relatively small sample sizes, unequal variances were assumed in independent-sample t-tests (i.e., Welch’s t-tests were used).

#### 2.3.2 Follow-up consultations

We additionally considered separately the requests for follow-up consultations, to gain insights into the medium-term outcome of the implementation of the program. Due to the low number of follow-up consultations, they could only be considered qualitatively.

## 3 Results

### 3.1. Consultations performed across participating institutions

The consultations analyzed took place from 10.07.2020 to 14.03.2021. Within the study period, 65 consultations were performed, four of which were follow-up consultations provided for patients who had already been examined previously, in order to follow the subsequent course of their condition and to potentially adapt diagnostics and therapy accordingly. The sample thus consists of 65 consultations for 61 patients. The following general evaluation of the data is based on the number of patients (N = 61).

Despite limitations imposed by the ongoing COVID-19 pandemic during the study period, the majority of consultations (N = 49, 80.3%) were performed on-site (Figure 1A). The number of requests varied considerably between the participating hospitals (N = 5; Figure 1B), significantly deviating from an equal distribution (*X*^2^ (4) = 77.93, p < .001), with more than half of the consultations requested by one hospital (Figure 1B).

**Figure 1:**
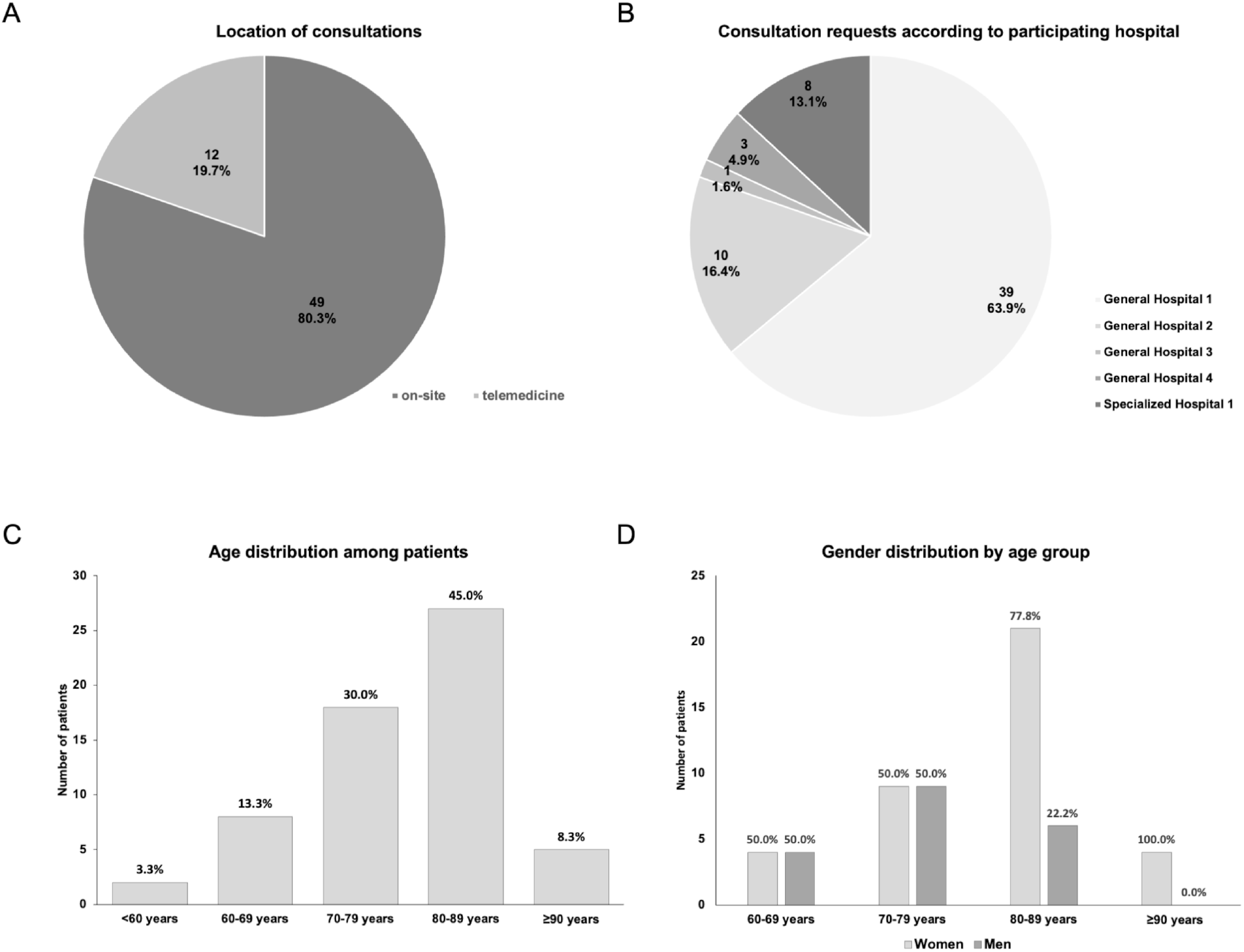
Basic conditions of the consultation service. A: Proportions of consultations performed on-site or via telemedicine. B: Distribution of the consultation requests among the participating hospitals. C: Age distribution among the patients for whom consultations were requested. D: Age distribution, separated by gender.

### 3.2. Demographic and clinical characteristics of the patients

#### 3.2.1. Demographics of the patients

Among the patients in our sample, more than half were 80 years or older (Figure 1C), and the number of women in the age groups of 80 years and above was higher compared to men (Figure 1D). The average age of women among the patients was 82.5 years (men: 73.1 years, t (27.7) = 2.94, p = .007). While this asymmetry was generally in line with the higher life expectancy among women, it should be noted that the proportion of men (n = 19) was still lower than would be expected based on the general population (2021).

#### 3.2.2. Conditions leading to hospital admission and indications for consultation requests

The conditions leading to acute admission to the partner hospitals were categorized according to the relevant medical specialty or the specific reason for admission. In absolute numbers, the most common reasons for patients’ admission to the partner hospitals were problems classified under Surgery (Figure 2A), with falls being the most frequently reported event leading to admission (N = 15, 24.6%). Other common reasons were categorized under Internal Medicine, Neurology or Psychiatry, and planned adaptation or modification of pharmacotherapy. The latter arose particularly at the partner hospital specializing in the treatment of diabetes. Notably, in 17 cases (27.9%), the primary reasons for admission to the partner hospitals were neuropsychiatric problems, although the partner hospitals had neither neurology nor psychiatry departments.

**Figure 2:**
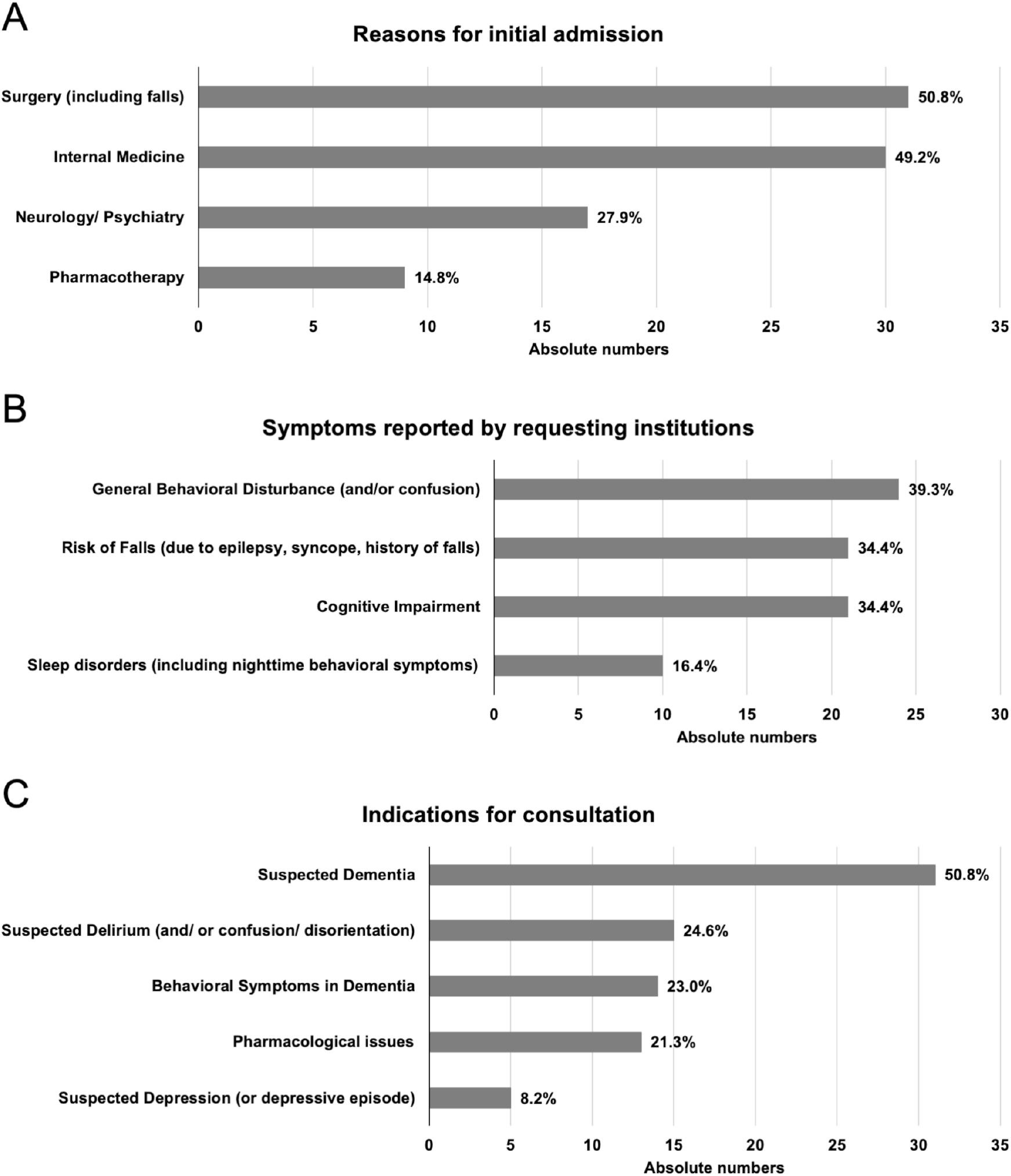
Clinical history and indications for consultations. A: Reasons leading to initial admission of the patients. B: Symptoms reported by requesting institutions. C: Specific indications for consultation. Total numbers sum to >100% as multiple items could be reported for one patient.

The most common symptoms reported to the service by the partner institutions when requesting a consultation could be classified as being related to *Behavioral Symptoms, Risk of Falls, Cognitive Impairment*, and *Sleep Disorders* (Figure 2B). Following data extraction, the questions were classified as indications for consultation and assigned to the following categories: *Suspected Dementia, Suspected Delirium, Behavioral Symptoms in Dementia, Pharmacological Issues* and *Suspected Depression* (Figure 2B).

In line with the focus of our consultation service on dementia, over half of consultation requests (50.8%, Figure 2C) were related to suspected dementia and/or cognitive impairment, and over a fifth (23.0%) specifically concerned behavioral symptoms in dementia. Questions relating to pharmacotherapy comprised both suspected adverse effects and requests for recommendations regarding pharmacological treatment.

#### 3.2.3. Pre-existing conditions and risk factors

A pre-existing confirmed diagnosis of dementia was reported in only nine consultation requests (14.8%) (Figure 3A). More broadly, a previous history of cognitive deficits was reported in 22 patients (36.1%), suggesting that dementia or cognitive impairment in older adults may be underdiagnosed among patients in rural primary care hospitals.

**Figure 3:**
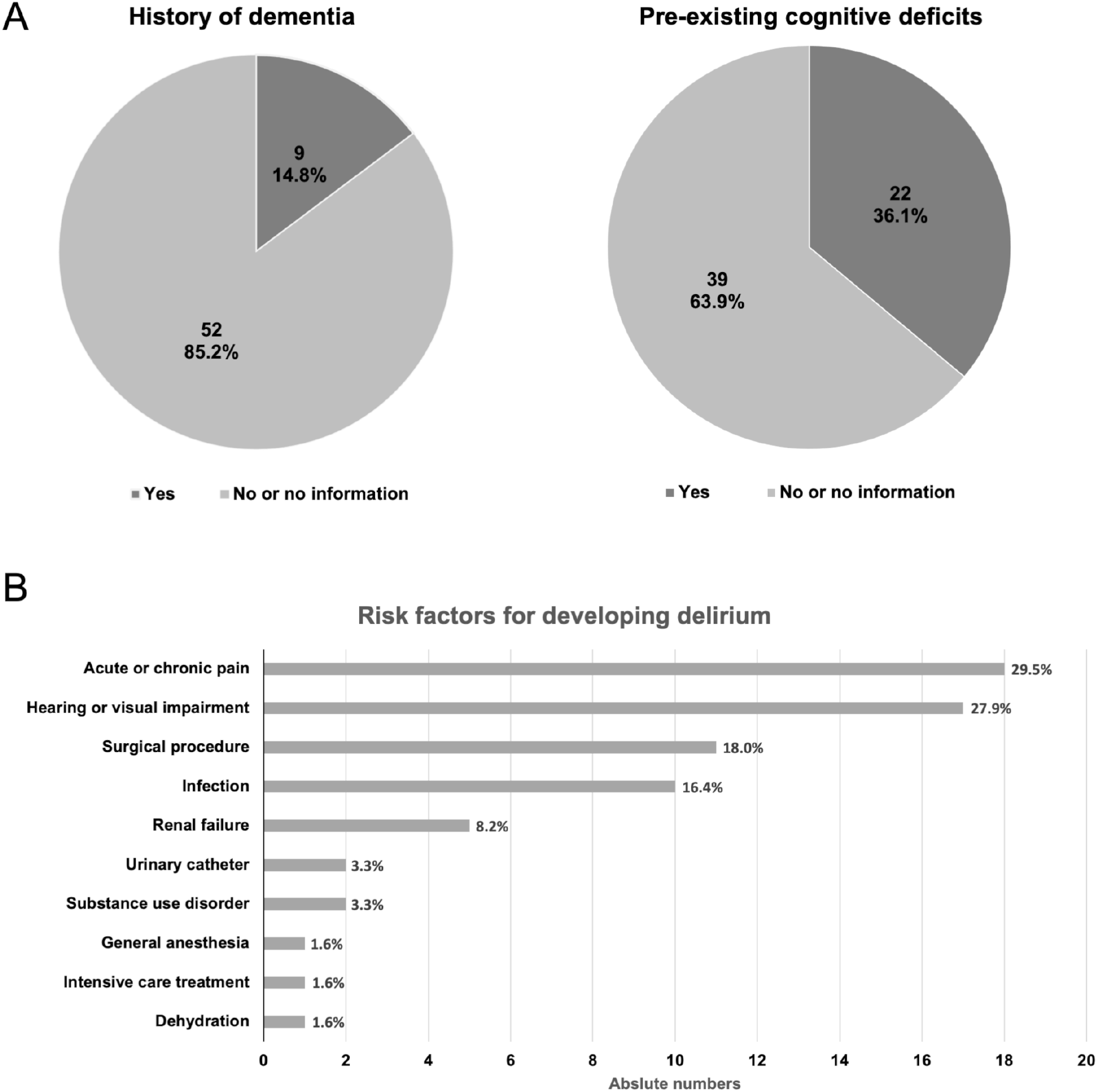
Clinical history related to dementia and risk for delirium. A: Proportions of patients with recorded history of dementia (left) or cognitive deficits (right), respectively. B: Risk factors for developing delirium. Total numbers sum to >100% as multiple items could be reported for one patient.

Beyond dementia and cognitive dysfunction, 38 patients (64.4%) had a history of at least three chronic medical conditions, and 23 (39.0%) had six or more diagnoses of a chronic disease. Considering the importance of delirium in the differential diagnosis for dementia and as a risk factor for morbidity and mortality in older adults (Inouye et al., 2014a), risk factors for developing delirium were specifically explored. Acute or chronic pain, hearing or visual impairment, recent surgical procedures, and infections were noted most frequently (Figure 3B).

### 3.3. Consultations

#### 3.3.1. Diagnostic assessment

Mirroring the focus of the consultation requests on suspected dementia, the group of dementias (ICD-10: F00 – F03) constituted the diagnoses made most frequently by the physicians performing the consultations (Figure 4A). Further diagnoses typically associated with cognitive symptoms included delirium (ICD-10: F05.x, F1x.4) and mild cognitive impairment (MCI; ICD-10: F06.7), a risk state for dementia (Schmidtke and Hermeneit, 2008). Notably, among the dementia subtypes, vascular dementias were most frequently recorded as diagnoses (Figure 4B), despite dementia in Alzheimer’s disease being the most common form of dementia in the population (2016). The MMSE (Folstein et al., 1975) was performed in 40 patients (Figure 4C), and the average MMSE score was lower in patients with a diagnosis of dementia (18.86 ± 5.16) than in those with other diagnoses (22.75 ± 5.01; t (21.7) = 22.23, p = 0.016). As expected, polypharmacy was highly prevalent among the patients referred for consultation, with a median number of 10 different medications (range: 2 – 23; mean = 10.31 ± 5.01; Figure 4D). There was, however, no difference in the number of medications as a function of dementia diagnosis or delirium (all p >0.468).

**Figure 4:**
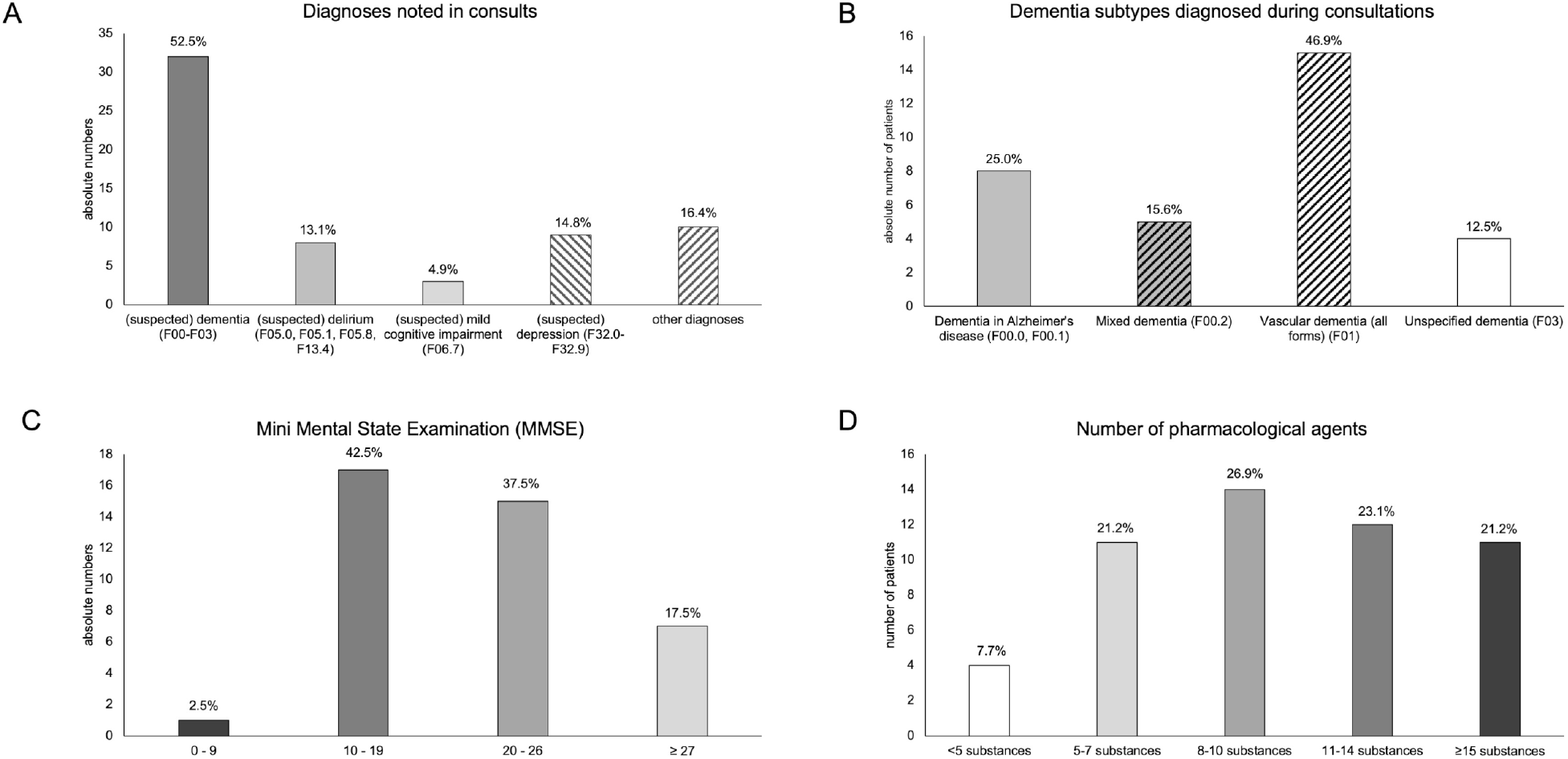
Diagnostic assessment during consultations. A: Primary diagnoses noted by the consulting physicians. B: Proportions of dementia subtypes among patients diagnosed with any form of dementia. Medium grey denotes the (suspected) presence of Alzheimer’s pathology. Hatch fill denotes (suspected) vascular pathology. C: MMSE scores among the patients investigated during consultations. D: Number of pharmacological agents taken by the patients.

#### 3.3.2. Recommendations

The physicians performing the consultations made a wide range of recommendations (Figure 5A). Recommendations regarding pharmacotherapy given most frequently (54.1% of patients). Among these, initiation of novel pharmacological treatment, including medication to be administered as required (*pro re nata*, PRN), was most commonly recommended, followed by discontinuation or reduction, and change of medication, which were recommended with equal frequency (Figure 5B). Non-pharmacological measures to treat behavioral symptoms were recommended for six patients (9.8%), but it should be noted that assistance with non-pharmacological interventions was primarily provided by the geriatric nurse (C.B.), who interacted with the nursing staff of the participating hospitals largely independently and provided advanced training in non-pharmacological treatment of dementia patients.

**Figure 5:**
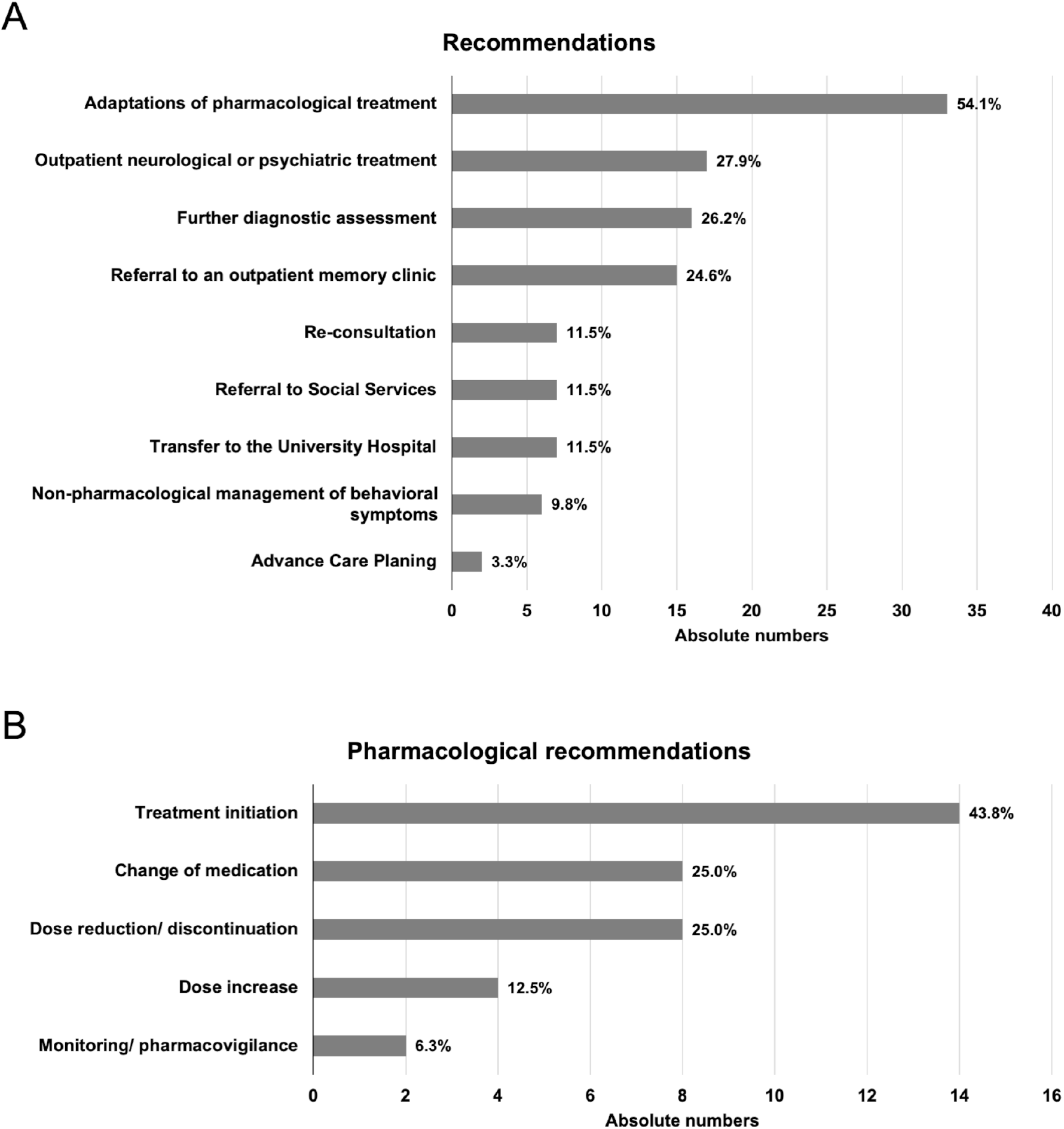
Therapeutic recommendations. A: General recommendations. B: Pharmacological recommendations. Total numbers add to > 100% as multiple recommendations could be given for one patient.

Further diagnostic assessment, referral for outpatient neuropsychiatric treatment, or consultation at a memory clinic were also frequently recommended. For seven patients (11.5%), transfer to a geriatric psychiatric inpatient ward was recommended. Follow-up consultations were recommended for seven patients but were subsequently requested and performed for four patients only.

#### 3.3.3. Follow-up consultations

Follow-up consultations were requested for three of the patients, with one patient receiving two follow-up visits. In two cases, follow-up visits were requested for patients who had initially been seen by the consultation service for suspected dementia and/or post-stroke depression after the patients had shown nocturnal episodes of confusion suggestive of delirium.

## 4 Discussion

The main goal of the study, the establishment of a combined outreach and telemedical dementia consultation service for rurally-based regional district general hospitals, was successfully achieved despite the obstacles posed by the ongoing COVID-19 pandemic. The requests received from the participating hospitals emphasize the need for the dissemination of expertise in dementia treatment and care among smaller hospitals in rural areas.

### 4.1. Implementation and evaluation of the consultations

The basic goal of the project, to improve the general care of people with dementia in rurally-based regional hospitals, was achieved: Agreements were signed with five partner hospitals, and consultations were performed for all five institutions, albeit with considerably varying frequency. Notably, over half of the consultations were requested by one of the participating hospitals. This tendency is probably best explained by the fact that this hospital has a geriatric department, which treats a large number of people with dementia, and the staff in this department may be more sensitive to the presence of cognitive or behavioral symptoms.

Across all participating hospitals, consultations were predominantly requested in relation to previously diagnosed or suspected dementia or associated behavioral problems (Figure 2C), confirming the need for a dementia-specific consultation service. Other frequent diagnoses included delirium, which is often difficult to distinguish cross-sectionally from dementia (Inouye et al., 2014b), and depressive syndromes, which are an important part of the differential diagnosis, especially in individuals with subjectively reported cognitive symptoms (Arie, 1983). We note the relatively low number of consultation requests (14.8 %) for patients with a pre-existing confirmed diagnosis of dementia. Although not explicitly stated in the information provided to the referring hospitals introducing the consultation service, our findings suggest that the service was considered to be useful primarily for establishing a new diagnosis, whereas patients who had previously received a diagnosis were likely to be already under regular medical care. Indeed, the predominant reason for requesting a consultation was for diagnosis, with suspected dementia (50.8 %), suspected delirium (24.6 %), and suspected depression (8.2 %) accounting for over three-quarters of the consultation indications.

Somewhat unexpectedly, the diagnosis of vascular dementia was the most frequently diagnosed form of dementia during the consultations. Given that dementia due to Alzheimer’s disease is the most common form in older adults (2016), we cannot exclude the possibility that some patients diagnosed with vascular dementia may nevertheless have had amyloid pathology and would thus have to be correctly classified as having mixed dementia. This differential diagnosis was, however, difficult to make as part of the consultation and should be performed, for example, in a memory clinic (Wallin et al., 2016). Alternatively, or more likely additionally, the proportionately high prevalence of vascular dementia in this patient group might reflect local risk profiles. The risk of major cardiovascular disease in Lower Saxony is higher than the average in Germany (Dornquast, 2016). Interestingly, however, a study previously conducted in the same region identified an under-diagnosis of Alzheimer’s dementia, with a corresponding over-diagnosis of an underlying vascular cause for dementia symptoms (Stoppe et al., 1994). Although a subsequent study found a reduction in the effect over the intervening years, the tendency nonetheless persisted (Maeck et al., 2007), which may contribute to the proportions among the diagnoses in our cohort.

More than 70% of the patients treated were receiving eight or more medications (Figure 4D), emphasizing the importance of considering the risks associated with polypharmacy in older patients, and particularly those with dementia, or more generally, cognitive impairment (Inouye et al., 2014a; Blair et al., 2018). Accordingly, in more than 50% of the consultations, an adjustment of pharmacotherapy was recommended, which commonly involved dose reduction or discontinuation, as well as replacement with medication with a more favorable side-effect profile (Figure 5). We note that the number of medications taken by a given patient was independent of patient age, degree of cognitive impairment, and diagnosis reached. Although such dependencies may become apparent in a larger study population, the finding underlines the importance of service provision for this patient group that is not limited to tight criteria but rather is based on communication with colleagues at the partner hospitals.

A direct transfer to a (geriatric) psychiatric inpatient unit was recommended in only 11.5% of cases. This low referral rate indicates that acute transfers of people with dementia to psychiatric departments – often associated with great distress for those affected – can possibly be at least partially avoided by providing adequate outreach consultation services. While further neurological or psychiatric assessment was indeed frequently recommended, it can often take place in an outpatient setting. The frequent recommendation of further diagnostic assessment, referral for outpatient neuropsychiatric treatment, or consultation at a memory clinic reflect the fact that dementia and cognitive impairment are chronic conditions that cannot be adequately treated in a single hospital stay. Access to the proposed consultation service meant that these unmet needs could be identified and seamlessly addressed by appropriate specialist facilities following the acute hospital stay, as opposed to requiring a subsequent separate assessment in the community before the necessary referral could be made.

The follow-up findings support the notion that the occurrence of delirium in patients with pre-diagnosed dementia is a common and clinically relevant complication warranting appropriate treatment. While the low number of follow-up consultations does not allow us to generalize from this observation, it is in line with the frequent occurrence of delirium in hospitalized patients with pre-existing dementia (Han et al., 2022).

### 4.2. Partnership with participating hospitals

Implementing a partnership with external hospitals (partly under private ownership) requires establishment of continuous communication. Individual approaches and intense exchange were essential in building a system of shared patient care. Gaining the interest of potential partners in the project was best achieved through personal contact, preferably on-site. Establishing contact via letter or e-mail remained more challenging, presumably due to the limited time capacities of the respective chief and senior physicians. Once the partnership had been established, it was essential to have fixed contact persons at the respective partner hospitals, who were aware of the possibility of dementia-specific consultations and could both request consultations and distribute information regarding our service. The establishment of a routine with fixed appointments and contact persons proved helpful for regular consultation requests. Telemedical consultations were not possible at all sites due to limited Wi-Fi and other technical factors. The successful implementation of this service in 12 cases, however, provides support for the feasibility of such an approach, which may grow in relevance during the current COVID-19 pandemic as well as future potential pandemics, in which this patient group is at high risk of serious consequences of infection, due to both advanced age and comorbidity. A specialist consultation service can provide diagnostic assessment and treatment plans without increasing social contact and infection risk.

The role of the nurse in providing non-pharmacological interventions underlines the requirement for a multidisciplinary service to address the clinical needs of this patient group, as highlighted in previous studies (Blair et al., 2018; Burton et al., 2021).

### 4.3. Limitations

A full evaluation of this consultation service requires a systematic, in-depth, long-term follow-up study. Future work should include a comprehensive quantitative and qualitative assessment of the impact of the implementation of the service. We suggest that the feasibility of the program implementation, as demonstrated here, provides the basis for such future work.

As only three patients received repeated visits, we could not derive quantitative information regarding the potential impact of the program on the longitudinal course of individual patients’ treatment. In a larger-scale future study, a greater number of patients would be expected to receive repeated consultations, which would enable collection of more representative information on medium-as well as long-term outcomes.

### 4.4. Conclusion and perspectives

Our findings indicate that provision of a dementia-specific consultation service for rurally-based, regional district general hospitals is feasible for university-based geriatric psychiatric units. We suggest that the concept of the dementia consultation service provided by a tertiary referral center (e.g., university hospital) to surrounding regional hospitals may provide a helpful addition to improve care for patients with dementia in rural areas. The service has the potential to reduce acute transfers to in-patient geriatric psychiatry. It enables older patients with dementia or delirium to be treated locally by assisting and empowering rurally-based regional hospitals to manage these problems and associated complications. Future studies are required to provide a more extensive evaluation of these effects. As our project was tailored to meet the capacities and needs of the German healthcare system, future research should also investigate to what extent the concept is transferrable to other geographic regions with comparable organization of hospital care.

## Data Availability

All original data are available from the corresponding authors upon reasonable request.

## Data Availability

All original data are available from the corresponding authors upon reasonable request.

## 5 Conflict of Interest

The authors have no conflict of interest, financial or otherwise, to declare.

## 6 Author Contributions

B.H.S., J.W., and K.R. conceived and designed the study and acquired funding; A.K. coordinated the consultation service; T.K. and C.A.T. performed the medical consultations; C.B. performed the nursing consultations; B.H.S. and K.R. supervised consultations; J.C.V., J.L.A., and A.K. performed data curation; B.H.S., J.C.V., and M.B. analyzed the data; B.H.S. and J.C.V. wrote the first draft of the manuscript; C.M.S.R., B.H.S., M.B. and K.R. interpreted the analyses and edited the manuscript; all authors contributed to editing and critically revising the manuscript.

## 7 Funding

This work was supported by the State of Lower Saxony, Department of Social Affairs, Health and Equal Opportunities (funding reference: 3SL_G.Demenz-04-2019). B.H.S. holds a grant from the European Union and the State of Saxony-Anhalt (Research Alliance *Autonomy in Old Age*). The funders had no role in the design or conduction of the study.

## 8 Acknowledgments

The authors would like to thank the staff of all participating hospitals. We thank Thomas Voigt and Matthias Siegert from the legal department of the UMG for support drafting the agreements and Petra Arndt, Hermann Esselmann, Ute Gotthardt, Cornelia Hönicke, Tobias Urbanczyk, and David Zilles for help with project administration. We further thank Hermann Esselmann for helpful comments on this manuscript.

## 9 Ethics Statement

This work was approved by the Ethics Committee of the University Medical Center Göttingen (approval number 13_12_19). Informed consent to be examined and treated by a physician from the University Medical Center Göttingen (UMG) was obtained from the patients or their caregivers by the participating hospitals. Additional consent to participate in research was waived by the Ethics Committee of the UMG, as all consultations were performed according to the current S3 Guidelines Dementia, and all data were processed and analyzed in a fully anonymized way.

## 10 Data Availability Statement

Data from this study are available in anonymized form from the corresponding authors upon reasonable request.

## Notes

### Competing Interest Statement

The authors have declared no competing interest.

### Clinical Trial

DRKS00022366

### Clinical Protocols

https://www.drks.de/drks_web/navigate.do?navigationId=trial.HTML&TRIAL_ID=DRKS00022366

### Author Declarations

The study was approved by the Ethics Committee of the University Medical Center Göttingen (approval number 13/12/19) and was registered with the German Clinical Trials Register (registration number DRKS00022366).

### Summary of Updates

Revised manuscript, including qualitative evaluation of follow-up consultations and a Limitations section

## References

Alzheimer’s Association (2016). 2016 Alzheimer’s disease facts and figures. Alzheimers Dement 12, 459–509.

Niedersächsisches Ministerium für Soziales Gesundheit und Gleichstellung (2016) [Online]. Available: https://www.ms.niedersachsen.de/startseite/service_kontakt/presseinformationen/demenzkranke-im-krankenhaus-besser-versorgen-land-foerdert-modellprojekte-mit-15-millionen-euro-175936.html.

Statistisches Bundesamt (2021) [Online]. Available: https://www.destatis.de/DE/Themen/Querschnitt/Demografischer-Wandel/_inhalt.html.

Arie, T. (1983). Pseudodementia. Br Med J (Clin Res Ed) 286, 1301–1302.

Blair, A., Anderson, K., and Bateman, C. (2018). The “Golden Angels”: effects of trained volunteers on specialling and readmission rates for people with dementia and delirium in rural hospitals. Int Psychogeriatr 30, 1707–1716.

Burton, J.K., Craig, L.E., Yong, S.Q., Siddiqi, N., Teale, E.A., Woodhouse, R., Barugh, A.J., Shepherd, A.M., Brunton, A., Freeman, S.C., Sutton, A.J., and Quinn, T.J. (2021). Non-pharmacological interventions for preventing delirium in hospitalised non-ICU patients. Cochrane Database Syst Rev 7, Cd013307.

Cao, Q., Tan, C.C., Xu, W., Hu, H., Cao, X.P., Dong, Q., Tan, L., and Yu, J.T. (2020). The Prevalence of Dementia: A Systematic Review and Meta-Analysis. J Alzheimers Dis 73, 1157–1166.

Chenoweth, L., Williams, A., Fry, M., Endean, E., and Liu, Z. (2021). Outcomes of Person-centered Care for Persons with Dementia in the Acute Care Setting: A Pilot Study. Clin Gerontol, 1–15.

Dewing, J., and Dijk, S. (2016). What is the current state of care for older people with dementia in general hospitals? A literature review. Dementia (London) 15, 106–124.

Dornquast, C.K., Lars E.; Neuhauser, Hannelore K.; Willich, Stefan N.; Reinhold, Thomas Busch, Markus A. (2016). Regional differences in the prevalence of cardiovascular disease—results from the German Health Update (GEDA) from 2009–2012. Dtsch Arztebl Int 113, 704–711.

Folstein, M.F., Folstein, S.E., and Mchugh, P.R. (1975). “Mini-mental state”. A practical method for grading the cognitive state of patients for the clinician. J Psychiatr Res 12, 189–198.

Han, Q.Y.C., Rodrigues, N.G., Klainin-Yobas, P., Haugan, G., and Wu, X.V. (2022). Prevalence, Risk Factors, and Impact of Delirium on Hospitalized Older Adults With Dementia: A Systematic Review and Meta-Analysis. J Am Med Dir Assoc 23, 23-32.e27.

Inouye, S.K., Marcantonio, E.R., and Metzger, E.D. (2014a). Doing Damage in Delirium: The Hazards of Antipsychotic Treatment in Elderly Persons. Lancet Psychiatry 1, 312–315.

Inouye, S.K., Westendorp, R.G., and Saczynski, J.S. (2014b). Delirium in elderly people. Lancet 383, 911–922.

Maeck, L., Haak, S., Knoblauch, A., and Stoppe, G. (2007). Early diagnosis of dementia in primary care: a representative eight-year follow-up study in Lower Saxony, Germany. Int J Geriatr Psychiatry 22, 23–31.

Marvanova, M. (2016). Drug-induced cognitive impairment: Effect of cardiovascular agents. Ment Health Clin 6, 201–206.

Mathur, S., Walter, S., Grunwald, I.Q., Helwig, S.A., Lesmeister, M., and Fassbender, K. (2019). Improving Prehospital Stroke Services in Rural and Underserved Settings With Mobile Stroke Units. Front Neurol 10, 159.

Mayring, P., and Fenzl, T. (2014). “Qualitative Inhaltsanalyse,” in Handbuch Methoden der empirischen Sozialforschung, eds. N. Baur & J. Blasius. (Wiesbaden: Wiesbaden), 543–556.

Milbert, A., and Sturm, G. (2016). Binnenwanderungen in Deutschland zwischen 1975 und 2013. Informationen zur Raumentwicklung 2, 121–144.

Ní Chróinín, D., Francis, N., Wong, P., Kim, Y.D., Nham, S., and D’amours, S. (2021). Older trauma patients are at high risk of delirium, especially those with underlying dementia or baseline frailty. Trauma Surg Acute Care Open 6, e000639.

Park, S.K., Lim, T., Cho, H., Yoon, H.K., Lee, H.J., Lee, J.H., Yoo, S., Kim, J.T., and Kim, W.H. (2021). Comparative effectiveness of pharmacological interventions to prevent postoperative delirium: a network meta-analysis. Sci Rep 11, 11922.

Schmidtke, K., and Hermeneit, S. (2008). High rate of conversion to Alzheimer’s disease in a cohort of amnestic MCI patients. Int Psychogeriatr 20, 96–108.

Sica, D.A. (2004). Diuretic-related side effects: development and treatment. J Clin Hypertens (Greenwich) 6, 532–540.

Stoppe, G., Sandholzer, H., Staedt, J., Winter, S., Kiefer, J., Kochen, M.M., and Rüther, E. (1994). Diagnosis of dementia in primary care: results of a representative survey in lower Saxony, Germany. Eur Arch Psychiatry Clin Neurosci 244, 278–283.

Van Den Berg, N., Grabe, H.J., Freyberger, H.J., and Hoffmann, W. (2011). A telephone- and text-message based telemedical care concept for patients with mental health disorders--study protocol for a randomized, controlled study design. BMC Psychiatry 11, 30.

Wallin, A., Nordlund, A., Jonsson, M., Lind, K., Edman, Å., Göthlin, M., Stålhammar, J., Eckerström, M., Kern, S., Börjesson-Hanson, A., Carlsson, M., Olsson, E., Zetterberg, H., Blennow, K., Svensson, J., Öhrfelt, A., Bjerke, M., Rolstad, S., and Eckerström, C. (2016). The Gothenburg MCI study: Design and distribution of Alzheimer’s disease and subcortical vascular disease diagnoses from baseline to 6-year follow-up. J Cereb Blood Flow Metab 36, 114–131.

